# Medical overuse in the Iranian healthcare system: a systematic scoping review and practical recommendations for decreasing medical overuse during unexpected COVID-19 pandemic opportunity

**DOI:** 10.1101/2020.05.10.20088369

**Authors:** Mohammad Zakaria Pezeshki, Ali Janati, Morteza Arab-Zozani

**Affiliations:** Social Determinants of Health Research Center, Department of Community and Family Medicine, Tabriz Medical School, Tabriz University of Medical Sciences, Tabriz, Iran; Iranian Center of Excellence in Health Management, Department of Health Services Management, School of Management and Medical Informatics, Tabriz University of Medical Sciences, Tabriz, Iran; Social Determinants of Health Research Center, Birjand University of Medical Sciences, Birjand, Iran; Student Research Committee, Tabriz University of Medical Sciences, Tabriz, Iran

**Keywords:** Medical Overuse, Healthcare system, Iran

## Abstract

**Background:** Overuse of medical care is a major problem across health systems as well as in Iran. By our knowledge, this is the first scoping review in which medical overuse in the Iranian healthcare system was investigated.

**Objective:** To perform an inclusive search for original studies that report medical overuse in the Iranian healthcare system.

**Methods:** A systematic search of the literature conducted in bibliographic databases including PubMed, Embase, Scopus, Web of Sciences, Cochrane and Scientific Information Database using a comprehensive search strategy without time limit until the end of 2018, accomplished by reference tracking, author contacting and expert consultation to identify studies on the overuse of medical care.

**Results:** We reviewed 4124 published articles based on predetermined inclusion criteria. The author’s consensus included a total of 40 articles. Of these, 31 were in English and 9 in Farsi, published between 19975-2018. The result categorized into two distinct clinical areas: treatment (17 articles), and diagnostic (23 articles) services. Almost all of the studies only described the magnitude of unnecessary overuse. Unnecessary overuse of Antibiotics, MRI, and CT-scan were the most reported topics. The ranges of their overuse proportion were as follows; Antibiotic (31 to 97%); MRI (33 to 88%), and CT-scan (19 to 50%).

**Conclusions:** Our review showed, even so, the magnitude of unnecessary overuse of medical services is high but there are only a few interventional studies in clinical and administrative level for finding effective methods for decreasing these unnecessary services. Researchers should be encouraged to conducting interventional studies. We suggest the ministry of health to use the golden opportunity of COVID-19 epidemic for designing Iran national policy and action plan for controlling and preventing unnecessary health care services and including a section for “Interventional Research” in the action plan.

## Introduction

Medical overuse means services that “are more harmful than beneficial, does not seem to increase the quality and quantity of life, impose excessive cost on the patients and their healthcare system, has low quality and if the patient has enough information, he or she will not ask for it” (1, 2). Overuse can take place in different area including medication, test or procedure (3). Recent studies have shown that overuse of tests and treatments can lead to serious consequences on patients in six domain including physical, psychological, social, financial, treatment burden, and dissatisfaction with health care (4). Medical overuse can delay access to the goals of health systems-improved health, responsiveness, financial risk protection and efficiency-by increasing cost and decreasing quality of medical care (5).

Given the fact that medical overuse is one of the problems that can make system performances problematic, its identification is of great importance for a health system (6). In addition, overuse in medical care is one of the obstacles to achieving universal health coverage (UHC) (7). If we want to achieve better UHC, we need to be able to manage costs, and one of the most important tasks in this direction is to reduce overuse in medical services (8, 9).

Over the years, many efforts have been made to identify overuse in medical care across health systems around the world (10, 11). In Iran, as in many other countries, there is little evidence about the amount of medical overuse in the health care system (1). So, the identification of medical overuse is an essential issue for the Iranian healthcare system and also helps health policymakers, health and medical managers, researchers, general practitioners, patients and their families to cope with harms, costs and quality of services. Ultimately, identifying overuse of medical care can bring our health system to balance in the right use of services. Thus, the objectives of this systematic scoping review were 1) to review the literature on the overuse of medical care, (2) to identify the areas in which the overuse of medical care take placed (3) to determine the rate of overuse of medical care in the Iranian healthcare system and its drivers and (4) to identify the interventional studies in clinical and administrative level for decreasing the rate of overuse

## Methods

We conducted a systematic scoping review on medical overuse in accordance with the five stages outlined in the Arksey and O’Malley framework (12). The review protocol was registered in PROSPERO before starting our study (registration no. CRD42017075481).

### Stage 1: Identifying research questions

The following questions guided this scoping review of medical overuse in the Iranian healthcare system: How many literatures have dealt with this issue in the Iranian healthcare system? Which area does medical overuse occur? What is the rate of medical overuse in the Iranian healthcare system? What are the drivers and preventive ways of medical overuse in the Iranian healthcare system?

### Stage 2: Identifying relevant studies

All original articles that investigated the overuse in medical care were included in the study. Of these, only studies were included that has addressed overuse in the Iranian healthcare system. All the included studies were limited in English and Farsi languages. Articles were excluded if the researchers did not have access to the Full-text.

We searched six databases including PubMed, Web of Science, Embase, Scopus, Cochrane and Scientific Information Database (SID) without time limit until the end of 2018. We also contacted the authors of included studies and use reference tracking to get the articles we probably did not find in the search. We used a set of Medical Subject Headings (MeSH) terms and free term to maximize the sensitivity of the search. For more information on search strategy, see study protocol at: https://bmjopen.bmj.com/content/8/4/e020355#ref-17 (1).

### Stages 3 and 4: Study selection and data charting process

After the search was completed, duplicate records were removed. Then, two reviewer screened the records based on title, abstract and full text, and extracted the data about authors, publication year, type of study, study population, type of service, clinical area, and overuse rate or range. These set of checklists designed for various types of studies and are available online (http://joannabriggs.org/research/critical-appraisal-tools.html). All potential disagreements in each level of study were resolved by consensus with a third researcher.

### Stage 5: Summarizing results

We categorized the results of the included studies based on publication year, clinical area, type of service (diagnostic tests, therapeutic procedures and medications), and range or rate of overuse.

## Results

Initially, a total of 4179 records were screened. After removing duplicates, 3023 records were considered for eligibility. Of these, 40 studies were included. The study selection process is outlined in the Preferred Reporting Items for Systematic Reviews and Meta-analysis (PRISMA) diagram (Figure 1).

**Figure 1:**
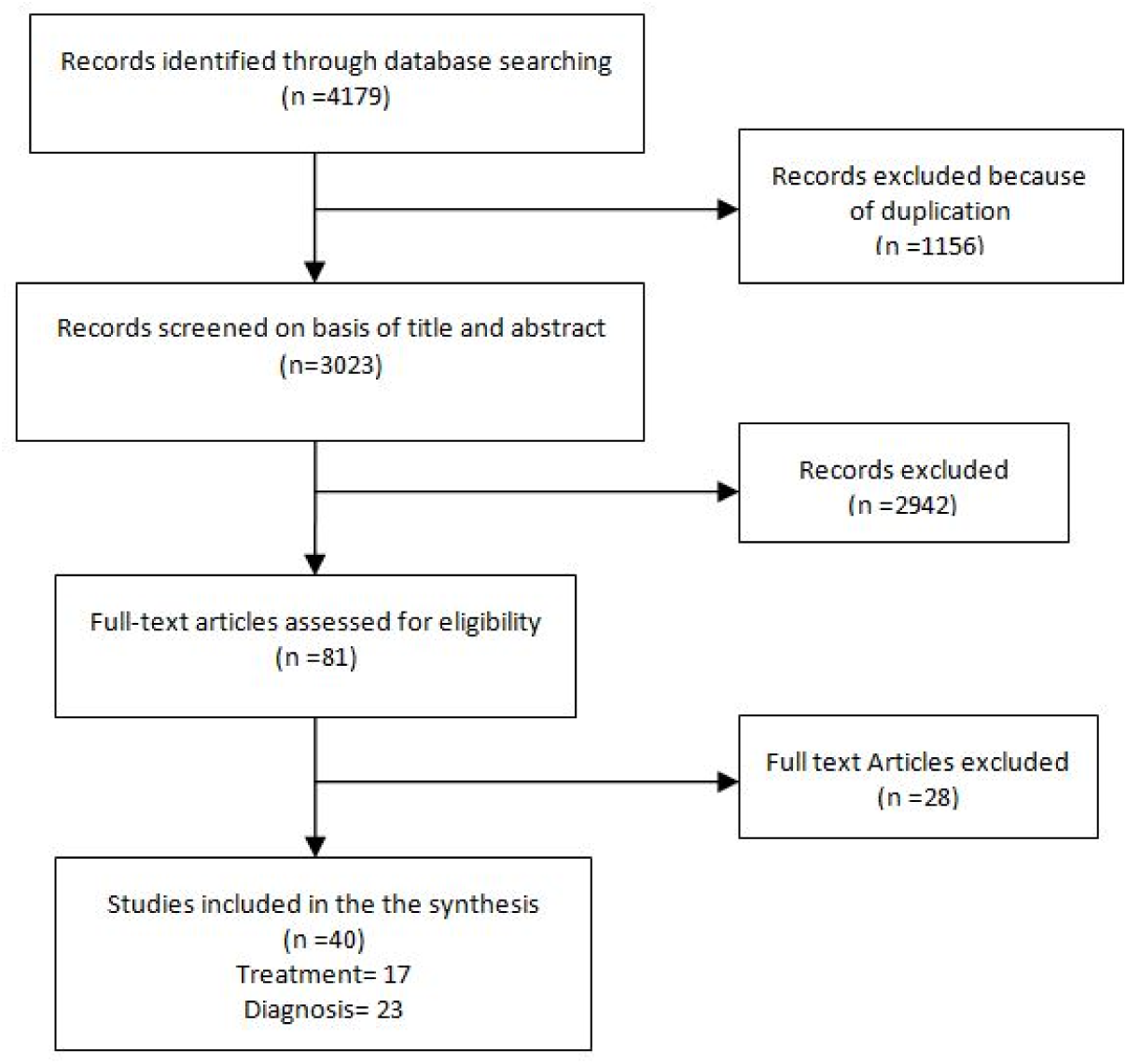
study flow diagram

Most studies were published in English (77.5%). Included studies published between 1975 and 2018. Most studies were published in 2014 (25%), 2012 (15%) and 2011 (12.5%). Also, in term of design, 37 were cross-sectional, and three RCT studies. The summary characteristics of included studies are shown in Tables 1.

**Table 1:**
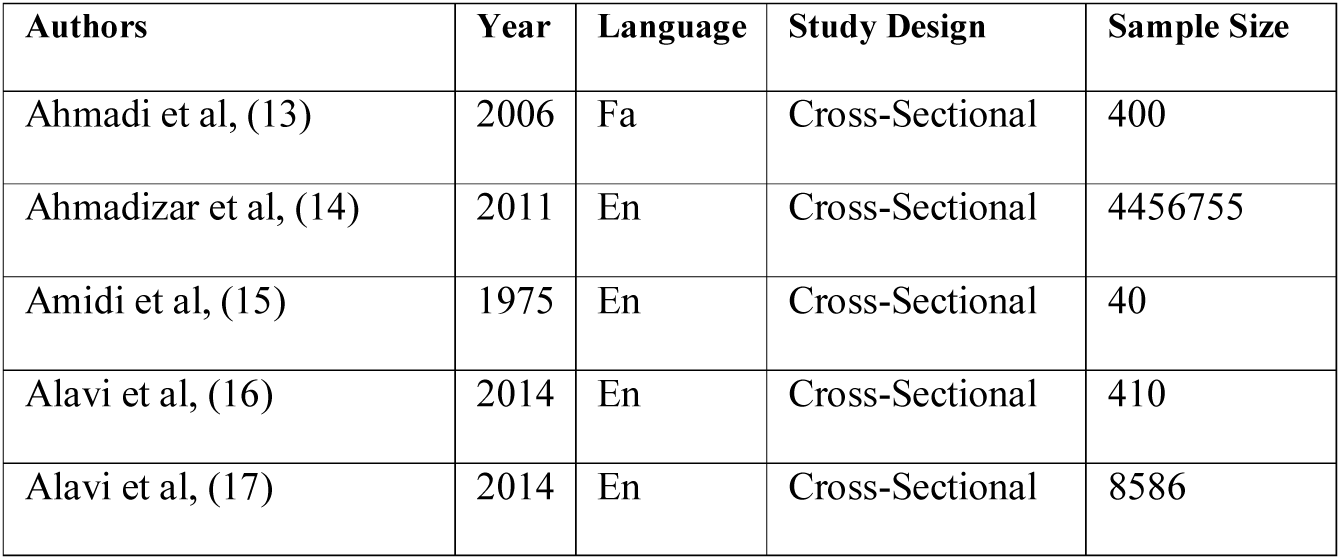
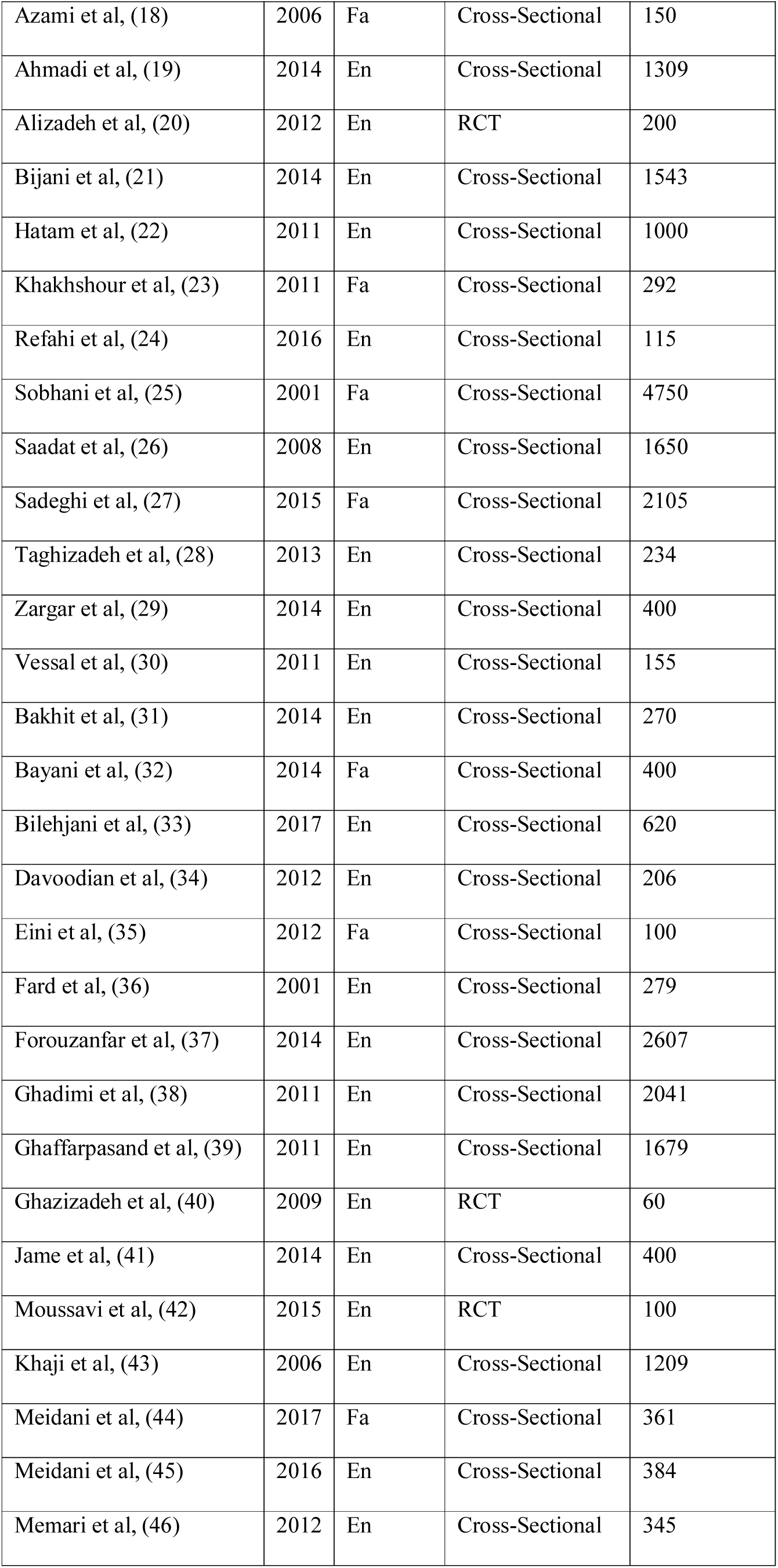
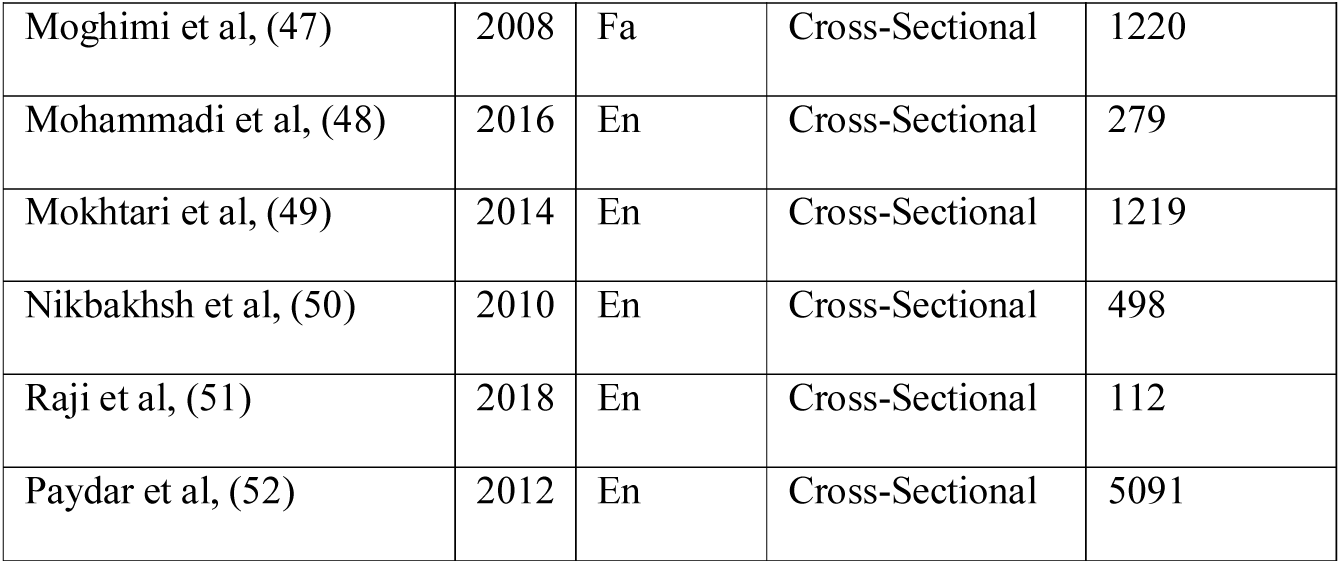
The summary characteristics of the included studies

The result categorized onto two distinct clinical area: treatment (17 articles), and diagnostic (23 articles) area. Unnecessary overuse of Antibiotics, MRI and CT-scan were the most reported topics. The ranges of their overuse proportion were as follow: Antibiotic (31 to 97%); MRI (33 to 88%), and CT-scan (19 to 50%). Among the studies in the area of treatment, the most studied were antibiotics (7 studies, 41.1%), and three studies (17.6%) did not compare the results with any other standard. Also, among the studies in the area of diagnosis, the most studied were related to MRI (4 studies, 17.3%), and CT (4 studies, 17.3%), and three studies (13%) did not compare the results with any other standard. For more details see Table 2 and 3.

**Table 2:**
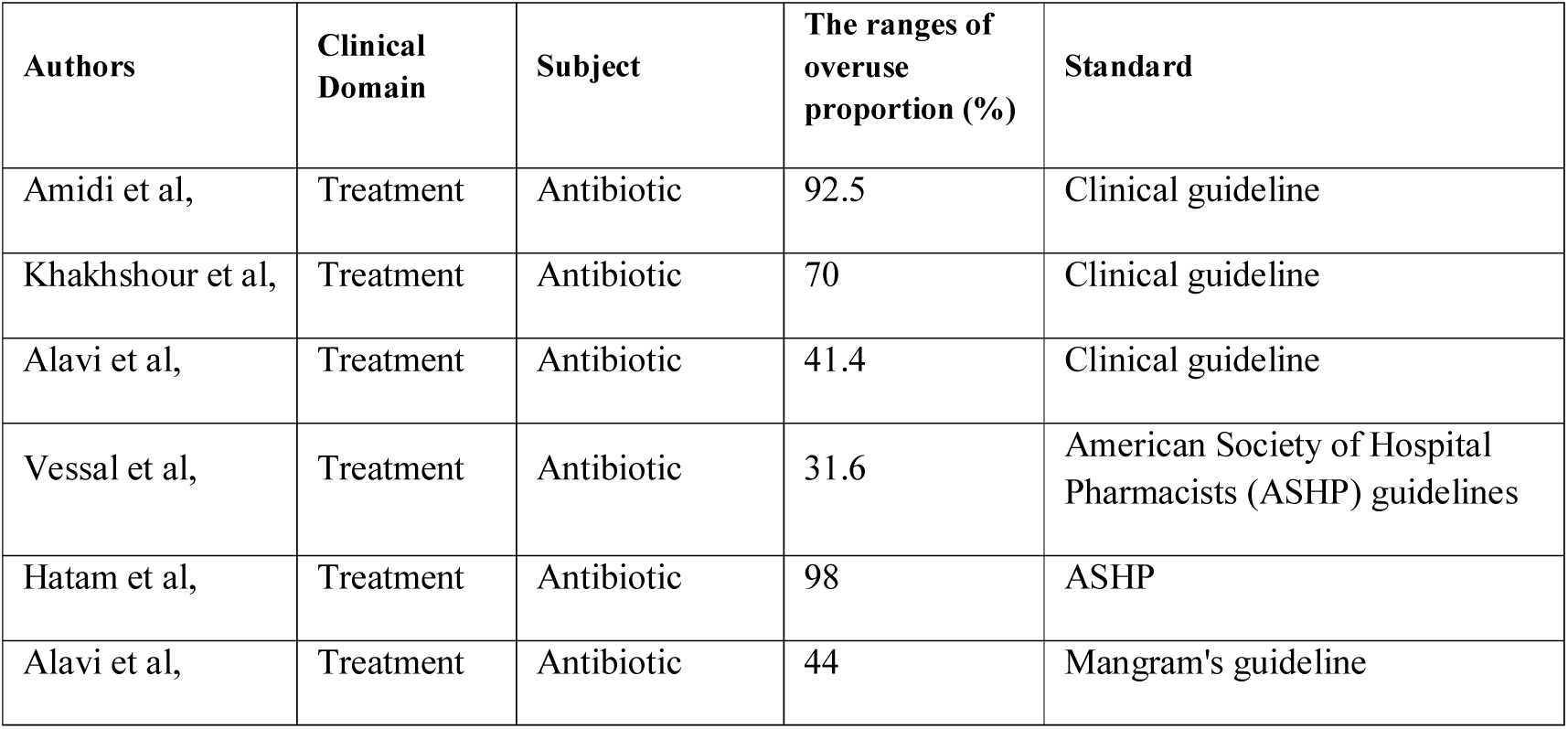
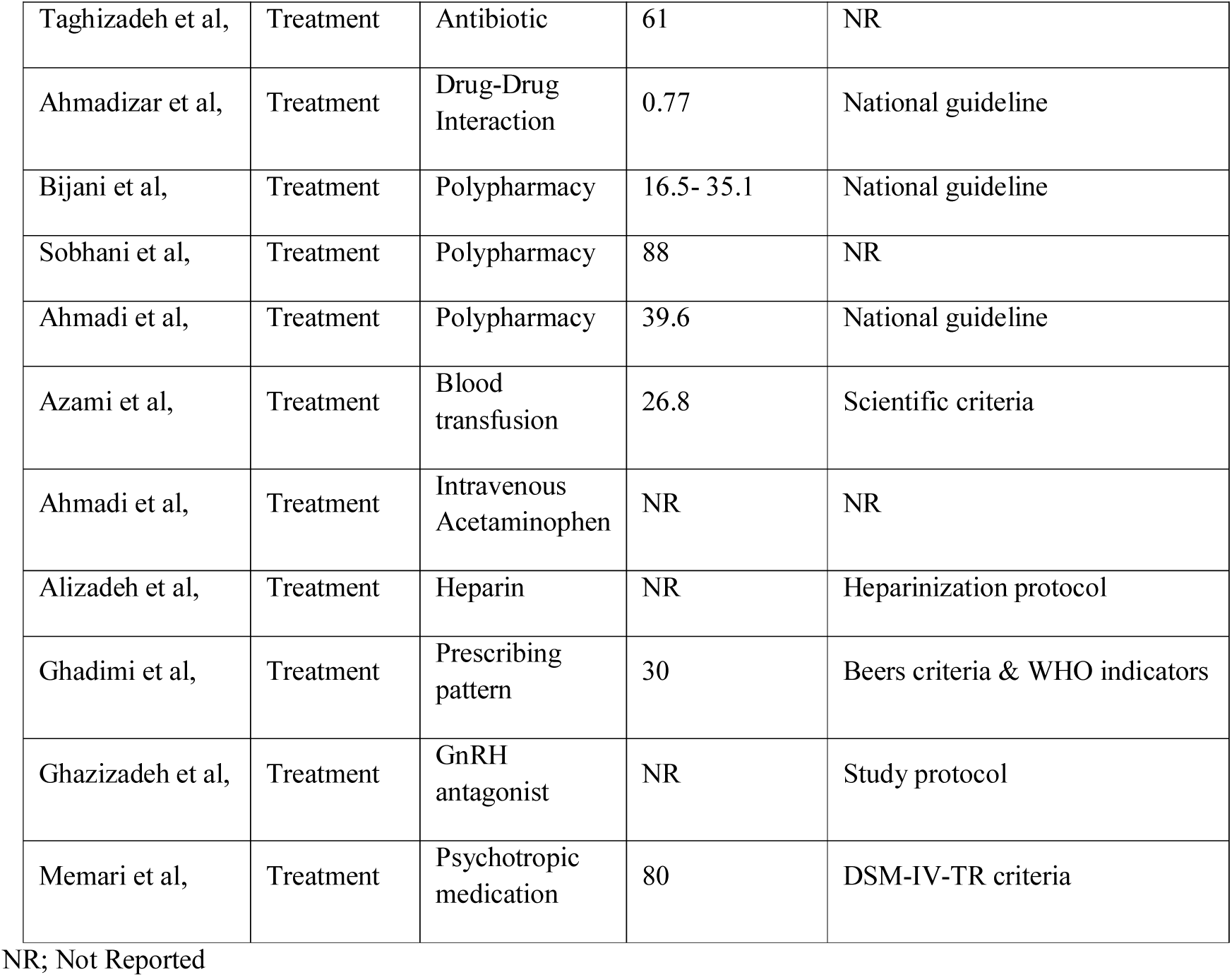
The characteristics of included studies in treatment area

**Table 3:** The characteristics of included studies in diagnostic area

| Authors | Clinical Domain | Subject | The ranges of overuse proportion (%) | Standard |
| --- | --- | --- | --- | --- |
| Refahi et al, | Diagnostic | MRI | 45.2 | Local guideline |
| Zargar et al, | Diagnostic | MRI | 46.5 | Clinical guideline |
| Saadat et al, | Diagnostic | MRI | 82.8 | Clinical guideline |
| Sadeghi et al, | Diagnostic | MRI | 76 | Clinical guideline |
| Bakhit et al, | Diagnostic | Diagnosis of dizziness | NR | Clinical guideline |
| Bayani et al, | Diagnostic | Clinical diagnosis and candida culture | NR | Clinical guideline |
| Bilehjani et al, | Diagnostic | Erythrocyte Sedimentation Rate (ESR) | NR | NR |
| Davoodian et al, | Diagnostic | Urinary catheters | 20.6 | NR |
| Eini et al, | Diagnostic | Antibacterial therapy | 97 | Clinical guideline |
| Fard et al, | Diagnostic | Venous duplex ultrasonography (VDUS) | NR | Scientific criteria |
| Forouzanfar et al, | Diagnostic | Chest X-ray (CXR) | 7.5 | Thoracic Injury Rule out Criteria (TIRC) |
| Ghaffarpasand et al, | Diagnostic | Radiography | NR | ATLS protocol |
| Jame et al, | Diagnostic | Computed tomography | 19.8-51.6 | Glasgow coma score |
| Moussavi et al, | Diagnostic | Computed tomography | NR | Glasgow coma score |
| Khaji et al, | Diagnostic | Computed tomography | 66.5 | Glasgow coma score |
| Meidani et al, | Diagnostic | Computed tomography | 14.1 | ACR criteria |
| Meidani et al, | Diagnostic | Laboratory test | 26.4 | ACR criteria |
| Moghimi et al, | Diagnostic | Preclinical test | 1.3-9.6 | NR |
| Mohammadi et al, | Diagnostic | MRI | 33 | NICE and AHRQ guidelines |
| Mokhtari et al, | Diagnostic | Venous thromboembolism (VTE) prophylaxis | NR | ACCP guidelines |
| Nikbakhsh et al, | Diagnostic | electrocardiogram (ECG) | 77.3 | American Society of Anesthesiologists status (ASA) criteria |
| Rajier al, | Diagnostic | Pulmonary CT angiography | NR | Geneva score and Wells' criteria |
| Paydar et al, | Diagnostic | Routine chest radiography for stable blunt trauma | 19.8 | ATLS |

NR; Not Reported

## Discussion

The aim of this systematic scoping review was to perform an inclusive search for original studies that report medical overuse in the Iranian healthcare system. Finally 40 original studies were included in our study, of which 17 article related to treatment area and 23 article related to diagnostic area. Antibiotics and MRI were the most reported issues in each category where overuse has been reported.

As Table 2 and Table 3 shows the majority of studies only have focused on the magnitude of unnecessary diagnostic and treatment services. There are only few interventional studies regarding diagnostic and treatment services. Also there is not any study regarding unnecessary clinical preventive services like unnecessary check up and also unnecessary public health services. Unfortunately as the Tables shows there is not any study at regional or national level that clarify the drivers of unnecessary services in Iran and how to address them. To address the shortage of study regarding the interventions for decreasing overuse rate in Iran we already conducted a qualitative research at national level to clarify the drivers of overuse and strategies for controlling these drivers in Iran. In this qualitative study we did interview with 21 well respected old hand policy makers and researchers of Iran. After analyzing the interview, our study showed that the main drivers of unnecessary overuse in the Iranian healthcare system are physician, patient, organizational, socio-cultural, market, and mass media factors. Also, a Policy Delphi analysis as part of our national study and based on the key informant’s opinion (53, 54), showed that the main interventions for decreasing unnecessary overuse of medical services include; implementing strategic purchasing, active engaging of insurance companies, promoting payment system, use of clinical practice guideline in decision making, and increasing political commitment and reducing conflicts of interest. We are going to publish the results of our study in detail.

COVID-19 pandemic has created a golden opportunity for addressing the drivers of unnecessary overuse of medical services by countries because of the three main reasons: 1) There is shortage of health care resources for controlling COVID-19 pandemic and unnecessary services waste the resources 2) Overuses of health care services unnecessarily expose the patients and healthy individuals to the virus in outpatient clinics and hospitals, 3) Overuse of medications may suppress the immune response and predispose people to COVID-19 infection. Our preliminary search shows that COVID-19 pandemic has decreased the use of several clinical interventions in countries (55) for example screening tests (56), admission and hospitalization (57), and elective surgeries (58, 59). Considerable proportions of these clinical interventions are unnecessary. We suggest Iranian ministry of health to use the golden opportunity of COVOD-19 pandemic to develop national policy and action plans for controlling and preventing unnecessary health care services in Iran. These policies will facilitate the controlling of COVID-19 epidemic and preventing underuse of necessary services during COVID-19 epidemic and after the end of epidemic.

## Conclusion

Our systematic review shows even so the magnitude of unnecessary overuse of medical services is high but there are only few interventional studies in clinical and administrative level for finding effective methods for decreasing these unnecessary services. Researchers should be encouraged for conducting such researches. It is necessary to be included a section for “Interventional Research” in the action plans we suggest to the ministry of health for controlling and preventing unnecessary health care services in Iran.

## Data Availability

All data included in the manuscript

